# Improving Visceral Leishmaniasis Service Delivery In Somalia: An Exploratory Qualitative Study Informed By CFIR-ERIC Matching Tool

**DOI:** 10.1101/2025.10.27.25338921

**Authors:** Abdirahman Moallim Ibrahim, Lul Mohamud Mohamed, Anoop Khanna, Mohamed Mohamud Fuje, Liban Hassan Jimale, Rimaz Mohamed Dewelbait, Mohamed Abdirahman Omer

## Abstract

**Background:** Visceral leishmaniasis (VL) is a life-threatening parasitic disease endemic in Somalia, where its control is severely challenged by a fragile health system, ongoing conflict, and socioeconomic barriers. This study aimed to assess the availability, accessibility, and quality of VL services in Somalia and to develop evidence-based, context-specific strategies to improve service delivery.

**Methods:** A qualitative study was conducted across five major VL treatment sites. Data were collected through in-depth interviews (n=57) with health managers and providers, focus group discussions (n=2) with patients and caregivers, and observational assessments using an adapted WHO Service Availability and Readiness Assessment (SARA) tool. The Consolidated Framework for Implementation Research (CFIR) guided data collection and thematic analysis to identify key determinants of implementation. The CFIR-ERIC Matching Tool was then employed to translate these findings into actionable strategies.

**Results:** Facility readiness assessments uncovered critical gaps and disparities in infrastructure, diagnostics, and essential medical supplies. CFIR analysis identified major barriers across multiple domains: the low adaptability and high complexity of VL protocols to the local context; unsustainable reliance on external funding; frequent service disruptions due to emergencies and outbreaks; and pervasive sociocultural misconceptions about the disease. Enablers included strong partnerships and committed local leadership. Based on this analysis, a set of tailored implementation strategies was developed, focusing on: creating flexible service delivery models (e.g., mobile clinics); securing sustainable financing; strengthening clinical and community referral networks; instituting continuous training and support for healthcare workers; and proactively engaging patients and communities.

**Conclusion:** The effective delivery of VL services in Somalia is impeded by a complex array of intervention-specific, systemic, and contextual barriers. This study provides a comprehensive assessment of these challenges and, crucially, generates a set of targeted, practical strategies to address them. Implementing these evidence-based, context-specific recommendations is essential for strengthening VL care and advancing progress towards disease elimination in Somalia.

## INTRODUCTION

Visceral leishmaniasis, also known as kala-azar, is a severe and often fatal parasitic disease caused by the protozoan parasite of leishmania genus and transmitted through the bites of infected sand fly [1,2]. It’s characterized by irregular fever, enlarged spleen and liver, anemia and weight loss [3–5]. It primarily affects the poor and most vulnerable population, particularly children and immunocompromised individuals [6]. The disease is endemic to parts of the world mainly in Southeast Asia and Eastern Africa with Somalia bearing a significant burden of the disease [5,7,8]. The context of Somalia is shaped by a combination of environmental, sociopolitical, and economic factors that worsen the public heath challenge posed by this disease [9–11].

Visceral leishmaniasis remains a major global public health problem in 4 eco-epidemiological regions of the world including the Americas, East Africa, North Africa, and West and South-East Asia [12]. Around 1 billion people live in areas endemic for visceral leishmaniasis and are at risk of contracting the disease. The global incidence of Visceral leishmaniasis is estimated to be 50,000-90,000 deaths annually [1]. In East Africa, visceral leishmaniasis is a significant public health problem with frequent outbreaks reported where sandflies are available [13,14]. The most prevalent countries include Sudan [15], South Sudan [16], Ethiopia [17], Kenya [18], Uganda [19] and Somalia [9]. The transmission dynamics of the disease are closely related to the presence of the vector in the arid and semi-arid regions of these countries. For instance, in Ethiopia, the disease is endemic in northwest lowland areas particularly in Amhara and Tigray region, and also prevalent in the Somali region across the border with Somalia [20–22]. In Kenya, the VL is endemic in the northwest and northeast regions, particularly Baringo, Turkana and NFD counties, where pastoralist communities are at high risk [23,24].

The region has experienced episodic outbreaks, often associated with displacement due to conflict, environmental changes, and the movement of non-immune individuals into endemic areas [25,26]. These outbreaks are further exacerbated by the limited access to healthcare services, diagnostic facilities, and effective treatment in remote and resource poor settings [27,28]. The parasites are maintained in a cycle involving sandflies and mammalian hosts including humans and rodents. The distribution of the disease is linked to the habitat preferences of the sandfly vectors and their interaction with human populations [29]. Changes in the environment including deforestation, urbanization, and irrigation can create favorable conditions for sandfly breeding and increase the risk of visceral leishmaniasis transmission [30].

Somalia, in particular, faces significant challenges in controlling and managing the disease [31]. Prolonged conflict and instability have fragmented and underfunded health services, destroying infrastructure and limiting access [32]. This creates difficulties in rural and conflict-affected areas for neglected tropical diseases like visceral leishmaniasis. In Somalia, the disease is predominant in semi-arid and arid southern regions, such as Lower Jubba, Bay, Bakool, and Gedo [9,33,34]. These areas suit sandfly vectors, and pastoralist communities there are at higher risk, often lacking healthcare, trained personnel, and supplies. Patients face barriers like long distances, insecurity, and financial constraints [9].

The complex interplay of these factors contributes to the persistence of and spread of the disease, particularly in areas with high levels of poverty and instability [28]. Therefore, understanding the challenges is crucial for developing control strategies tailored to the context and improving health outcomes for the effected population. The study aims to improve the delivery of visceral leishmaniasis services by assessing the current availability, accessibility and quality of VL services, identifying the existing barriers and enablers, and ultimately developing context-specific strategies and recommendations for effective service delivery.

## METHODS

### Study designs

The study employed a qualitative design to comprehensively assess visceral leishmaniasis service delivery and identify barriers and enablers. This included in-depth interviews, focus group discussion and observational assessments. Observational assessments were adapted from the World Health Organization’s Service Availability and Readiness Assessment (SARA) checklist to evaluate facility readiness, focusing on infrastructure, diagnostic tools, medical supplies, and overall capacity to provide visceral leishmaniasis services [35]. In-depth interviews and FGDs were guided by the Consolidated Framework for Implementation Research (CFIR) to explore implementation factors, including intervention characteristics, internal and external settings, individual roles, and processes [36]. The CFIR-ERIC Matching Tool was used to develop context-specific strategies that address identified CFIR barriers while leveraging enablers, ensuring recommendations were tailored to Somalia’s unique context [37]. The observational component involved site visits to assess infrastructure, diagnostic tools, medical supplies, and facility capacity, providing contextual insights into service delivery. In-depth interviews with healthcare providers, program managers, and decision-makers explored policy implementation, resource allocation, training, and external factors like conflict and displacement. Focus group discussions with patients and families offered grassroots perspectives on experiences, challenges, awareness, accessibility, quality of care, and socio-economic or cultural barriers.

### Study setting

The study was conducted across five significant sites in Somalia, chosen for their role in diagnosing and treating visceral leishmaniasis. Two of these sites are in the Bay Region: the Kala-azar Center, operated by SOS, which serves patients from Bay, Gedo, Middle Jubba, Bakool, and neighboring Ethiopian regions, and Bay Regional Hospital, a 200-bed referral hospital for southern Somalia and endemic areas. In the Benadir Region, three sites were included: SOS Hospital, providing primary care with a focus on pediatrics and treating suspected pediatric VL cases; De Martino Hospital, offering inpatient services for adult VL patients, and Benadir Hospital, a mother and child hospital that treats suspected pediatric VL cases. These facilities are key in providing comprehensive diagnostic and treatment services for VL in Somalia.

### Study participants and sampling procedure

The study employed purposive sampling to select participants who were most relevant to the objectives, ensuring the inclusion of individuals with direct knowledge and experience related to visceral leishmaniasis (VL) service delivery. The study included participants such as program managers, health service providers (including medical doctors, nurses, laboratory technicians, and pharmacists) and patients. A total of 57 in-depth interviews were conducted. These interviews were guided by the Consolidated Framework for Implementation Research (CFIR), ensuring a comprehensive exploration of all relevant aspects of VL service delivery. The interviews focused on key implementation elements, such as the characteristics of the VL interventions, and the influence of both internal (health system factors) and external (community and environmental) settings on service delivery.

In addition to the interviews, two focus group discussions (FGDs) were conducted, involving 16 participants, who were either patients with suspected VL or family members of affected individuals. These FGDs aimed to capture the perspectives and lived experiences of those directly impacted by VL, providing valuable insights into the barriers and enablers of VL service delivery from the patients’ point of view. The combination of interviews and FGDs helped to ensure a well-rounded understanding of the challenges and opportunities for improving VL services in Somalia.

### Researcher Characteristics and Reflexivity

The research team included three members with complementary expertise relevant to the study context. AMI, the principal investigator, is a public health researcher with extensive experience in tropical disease management and qualitative methods in Somalia. LMM, a research associate, brought specialized knowledge in health systems and community-based interventions in fragile settings. RMD, a methodology expert, contributed advanced skills in implementation science and qualitative analysis.

Given AMI’s previous engagements with health facilities in Somalia, specific reflexivity practices were implemented throughout the research process. AMI maintained a detailed reflexive journal to document preconceptions, emerging insights, and potential influences on data collection, particularly when interviewing healthcare providers at facilities where previous professional relationships existed. This journal served as a tool for critical self-reflection regarding positionality and power dynamics during interactions with various stakeholders - from ministry officials and healthcare providers to vulnerable patients and family members.

Weekly team discussions facilitated critical examination of how researchers’ backgrounds and perspectives might influence data interpretation. LMM’s background in health systems provided important contextual understanding, while RMD’s external perspective helped identify and challenge assumptions arising from familiarity with the setting. The team actively discussed how their professional identities and prior experiences might shape what they noticed, how they interpreted data, and the questions they asked during interviews and analysis. To mitigate potential biases, the team implemented several strategies: using multiple coders during analysis, seeking disconfirming evidence in the data, and explicitly discussing how their positions as educated health professionals might affect interactions with patients and frontline health workers. These practices enhanced the credibility of findings by ensuring that researchers remained attentive to how their positions and perspectives influenced the research process

### Implementation framework

The Consolidated Framework for Implementation Research (CFIR) 2.0 was used as the guiding framework to evaluate and improve visceral leishmaniasis (VL) service delivery in Somalia [38–40]. CFIR 2.0 is an updated version of the original CFIR framework, designed to provide a structured approach for assessing the implementation of interventions in complex healthcare settings. This comprehensive framework helps researchers and practitioners understand the myriad factors that can influence the success or failure of implementing new practices or improving existing ones. CFIR 2.0 organizes implementation factors into five major domains, each containing multiple constructs that represent different aspects of the implementation process: 1) Intervention characteristics, 2) Outer setting, 3) Inner setting, 4) Characteristics of individuals, and 5) Process. The aim of using CFIR 2.0 in this study was to systematically assess the various factors affecting the implementation of VL services in Somalia. The framework provided a comprehensive structure for identifying the barriers and enablers within each domain that could influence the success of VL service delivery. In addition to CFIR 2.0, the study utilized the CFIR-ERIC Matching Tool to develop and prioritize relevant strategies that address the identified barriers. The CFIR-ERIC tool is an evidence-based resource that matches specific CFIR-identified barriers to corresponding implementation strategies recommended by the Expert Recommendations for Implementing Change (ERIC) project [37,41–43].

### Data collection

Data were collected from March to July 2024 using a multi-method approach combining face-to-face and field visits. Site visits to the five VL treatment centers were conducted in person, allowing for comprehensive observational assessments using an adapted WHO Service Availability and Readiness Assessment (SARA) checklist. These on-site evaluations documented facility infrastructure, diagnostic capacity, medication availability, and overall service readiness through direct observation and inventory reviews.

Data collection included 57 in-depth interviews and 2 focus group discussions with 16 participants. Each interview lasted 45-60 minutes. All interviews were completed face-to-face in comfortable, private settings at health facilities or neutral community locations. The approach for each interview was determined based on participant preference and security considerations. Focus group discussions were conducted in person at neutral community locations near the treatment facilities to ensure participant comfort and accessibility. All qualitative interactions followed semi-structured guides developed using CFIR constructs, with the interview guide provided as supplementary file 1. With participant consent, all sessions were audio-recorded and transcribed verbatim. Data collection continued until saturation was reached, where no new themes emerged from additional interviews. All data collectors received specialized training on research ethics, interview techniques, and VL service context, and detailed field notes were maintained to capture non-verbal cues and contextual observations.

### Data analysis process

A convergent mixed-methods analytical approach was employed. Quantitative observational data from the SARA assessments were analyzed descriptively to calculate facility readiness scores across key domains including staffing, infrastructure, diagnostics, and essential commodities.

For qualitative data, thematic analysis was conducted using a deductive approach, with CFIR 2.0 as the primary coding framework. Audio recordings were transcribed verbatim by trained research assistants, with transcription accuracy verified by AMI through random checks of 30% of transcripts. All transcripts were anonymized by removing identifying information (names, specific organizations, exact locations) before analysis. Data were managed in Excel software, with each transcript assigned a unique identifier to maintain confidentiality throughout the analysis process. The coding process involved multiple stages. First, AMI and LMM independently coded five transcripts using the CFIR framework, meeting to resolve discrepancies and refine the coding structure. Once inter-coder reliability was established, the remaining transcripts were divided between researchers for independent coding. Regular team meetings were held to discuss emerging patterns and resolve coding disagreements through consensus, with RMD providing methodological oversight.

To enhance analytical rigor and trustworthiness, several verification strategies were employed. Analyst triangulation was achieved through independent coding and regular debriefing sessions. Member checking was conducted by sharing preliminary findings with eight participants (including health providers, program managers, and patients) to verify interpretive accuracy. A comprehensive audit trail was maintained to document all analytical decisions and theme development processes. Coded data were synthesized to identify prominent barriers and enablers across CFIR domains, with particular attention to interactions between different implementation factors. The research team then systematically applied the CFIR-ERIC Matching Tool to generate evidence-based implementation strategies, prioritizing those most feasible and relevant to the Somali context. The consolidated criteria for reporting qualitative research (COREQ) guided the reporting of all qualitative components of the study.

## ETHICAL CLEARANCE

Ethical clearance were obtained from the somali National Institute of Health (NIH) under the refference number **NIH/IRB/11/DEC/2023**. An informed consent was sought from all the participants in the study, ensuring they were fully informed and voluntarily participated in the research. For participants who were unable to provide written consent, verbal consent were obtained and documented appropriately.

## RESULTS

### STATUS OF VISCERAL LEISHMANIASIS CENTERS

From the observation field notes, the current access and quality of care for visceral leishmaniasis (VL) in Somalia were assessed across five key sites using the WHO Service Availability and Readiness Assessment (SARA) tool. The data collected from SOS Mogadishu, De Martino Hospital, Benaadir Hospital, SOS Baidoa, and Bay Regional Hospital revealed variations in the availability of essential services. The + signs in the table indicate the level of service availability, with "+" representing a lower grade of availability and "+++" representing the highest grade.

Staffing was consistently rated highly across all sites, indicating that most of these centers had adequate human resources to manage VL cases. However, infrastructure varied, with SOS Baidoa showing the highest level of infrastructure readiness, while other centers like De Martino Hospital and Bay Regional Hospital had relatively lower infrastructure scores. In terms of inpatient and observation beds, SOS Mogadishu, Benaadir Hospital, and SOS Baidoa performed better, while De Martino Hospital and Bay Regional Hospital had fewer beds available for patients.

Basic amenities such as water, sanitation, and electricity were mostly adequate, with all hospitals performing well in this category, except for Bay Regional Hospital, which showed slight limitations. The availability of medicines and commodities was also uneven, with De Martino Hospital and Bay Regional Hospital showing lower availability compared to other sites that maintained consistent supplies.

Diagnostics and information systems followed a similar pattern, with SOS Mogadishu and SOS Baidoa showing higher availability, while other hospitals lagged behind. This assessment highlights significant disparities in the availability of key resources necessary for effective VL care, underscoring the need for targeted improvements in certain facilities **(Table 1).**

**Table 1.** Status of visceral leishmaniasis centers.

### Barriers and enablers to improving visceral leishmaniasis service delivery

#### INTERVENTION CHARACTERISTICS

##### Adaptability of visceral leishmaniasis services

The adaptability of viscral leushmaniasis services emerged as a barrier in the effective implementation of VL diagnosis and treatment in the country. Many participants including program managers, health service providers, and patients expressed concerns about the current VL services particularly the treatment protocols in adapting to local context.

One program manager highlighted this issue, stating, "*The treatment protocols are very specific, and while they are based on strong evidence, they don’t always take into account the realities we face here in Somalia. For instance, we might not have all the recommended drugs available, or the diagnostic equipment might be outdated*."

A qualified nurse stated, "*We have to follow the protocols, but sometimes it’s impossible. The patients we see come from remote areas, and they can’t always return for follow-up visits. The guidelines don’t account for these kinds of challenges*." This illustrates how the lack of adaptability in the VL intervention can result in suboptimal care, particularly in regions where patients face significant barriers to accessing healthcare services. Patients and their families also felt the impact of the intervention’s limited adaptability. A family member of a patient expressed frustration, saying, "*We were told to come back for more tests, but we live far away, and it costs too much to travel. The doctors here are doing their best, but the system doesn’t work for us.*"

### The complexity of VL services

The protocols for diagnosing and treating VL are complicated, requiring specialized training, specific diagnostic tools, and a series of steps that must be followed precisely to ensure effective treatment. This complexity posed significant challenges for health service providers, particularly in settings with limited resources and infrastructure.

A program manager described the issue, saying, "*The VL treatment protocols are very detailed, which is good for ensuring quality care, but they are also very demanding. Not all of our staff are fully trained on every aspect, and the equipment we need isn’t always available*." This complexity was seen as a double-edged sword necessary for high-quality care but difficult to implement in practice.

Health service providers often struggled with the complexity of the intervention, particularly in terms of the diagnostic procedures. A laboratory technician shared, "*In the lab, we only use rapid tests to diagnose VL, while other preferred existing diagnostic tools including DAT and microscopic tests are not available and the lab staff are not even trained if they were available, so it’s very difficult to reach a final decision sometimes, even when the patient is clinically ill*."

### Cost of Implementing VL services

The cost of implementing VL services was frequently expressed as a barrier to effective service delivery. An exectutive manager explained the situation, stating, "*The cost of running these programs is very high. We rely heavily on support from international partners to fund everything from the medicines to the training programs. Without their help, we wouldn’t be able to provide these services at all."* This reliance on external funding highlights the vulnerability of the VL services in Somalia, as the sustainability of the intervention is closely tied to continued international support. Health service providers also expressed concerns about the financial constraints they faced. One doctor mentioned, "*We have to be very careful with our resources. Sometimes, we have to make difficult decisions about who can receive treatment based on what we can afford*." This underscores the harsh realities of working within a constrained budget, where financial limitations directly impact the quality and availability of care.

Patients, too, felt the impact of these financial constraints. A patient shared their experience, saying, "*The treatment is supposed to be free, but sometimes we are asked to pay for certain tests or medications because the hospital doesn’t have enough supplies*." This situation often led to delays in treatment or, in some cases, patients abandoning treatment altogether due to the costs involved.

### OUTER SETTING

#### Critical incidents: Emergencies and Outbreaks

The delivery of visceral leishmaniasis (VL) services in Somalia is deeply impacted by critical incidents, such as emergencies and disease outbreaks, which can severely disrupt the provision of healthcare. Many program managers, health service providers, and patients reported that these events often divert attention and resources away from VL services, leading to delays in diagnosis and treatment, and sometimes, the complete cessation of services.

A health manager described the challenges during the COVID-19 pandemic: "*When COVID hit, all our resources, including staff and supplies, were redirected to manage the emergency. VL services were almost entirely suspended, and patients had to wait longer for treatment, which is dangerous given the rapid progression of VL if untreated*."

Health service providers echoed this concern, noting the strain on the healthcare system during emergencies. A qualified nurse shared, "*When there’s an outbreak or any kind of emergency, we are overwhelmed. The focus shifts entirely to the emergency at hand, and VL patients are often left waiting. It’s heartbreaking because you know that these delays can be fatal*."

Patients and their families also experienced the consequences of these disruptions. A patient said, "*I was supposed to get my treatment, but when I came to the hospital, they told me they were dealing with an emergency and that I had to come back later. But I was already very sick and traveling back and forth is not easy."*

### Local attitudes: sociocultural values and beliefs

The participants noted that the misconceptions about the VL and its treatment are widespread, often leading to delays in seeking care or reluctance to adhere to treatment regimens.

A Health Manager stated *"There is a lot of misinformation about VL in the community. Some people believe that it’s caused by a curse or that traditional medicine is more effective than what we offer in the clinics. This makes it very difficult to get patients to come in for diagnosis and treatment."*

Health service providers also reported challenges related to local beliefs about VL. A doctor shared, *"Patients sometimes come to us only after they’ve tried traditional remedies that don’t work. By the time they arrive, the disease has often progressed to a more severe stage, making treatment more complicated and less effective."*

However, there were also examples of local attitudes serving as enablers. A health worker noted, *"In some communities, we’ve been able to work with traditional healers to refer patients to us. This has been very helpful because it means we’re catching cases earlier and can start treatment sooner."*

### Local conditions: Economic, environmental, political and technological factors

As participants highlighted, the economic situation, characterized by widespread poverty, limits the ability of many patients to access care, even when services are theoretically available.

A hospital director commented on the economic challenges: *"Many of our patients come from very poor backgrounds. Even though the treatment is free, the cost of transportation, time away from work, and other indirect costs make it difficult for them to access the services. This is a huge barrier."*

The participants also expressed that environmental conditions play a role, particularly in rural areas where access to healthcare facilities is limited by poor infrastructure and challenging terrain. A doctor noted, *"During the rainy season, roads become impassable, and patients simply can’t reach us. We’ve had cases where people couldn’t get treatment in time because they were stranded by floods."*

The political situation in Somalia, marked by instability and conflict, further complicated the delivery of VL services. An executive director explained, *"Insecurity in some regions makes it impossible to provide consistent services. Our staff can’t travel to certain areas, and we have to rely on local partners, who may not always have the capacity to deliver the same level of care."*

Technological limitations, such as the lack of advanced diagnostic tools and electronic health records, also hindered VL service delivery. A laboratory technician mentioned, *"We are still using very basic equipment, and there’s no way to share patient records between facilities. This makes follow-up difficult and leads to gaps in care."*

### Partnerships: referral networks, international and local partners

Many program managers and health service providers emphasized the importance of partnerships in overcoming the challenges associated with VL service delivery. A hospital director highlighted *"We have established strong referral networks with smaller clinics and community health workers. This means that even if we can’t reach every patient directly, we can ensure that they are referred to the nearest facility that can provide the necessary care."*

International and local partners also play a crucial role in supporting VL services, particularly in terms of funding, training, and providing essential supplies. An executive manager noted, *"Without the support of our international partners, we wouldn’t have the medicines, training, or even the basic equipment we need to treat VL. They have been instrumental in keeping these services running."*

### Policies and laws: Regulations, Guidelines, and the health system structure

Most of the participants noted that while there are guidelines in place, their implementation is often inconsistent due to a lack of oversight and resources.

A program manager explained, *"We have good guidelines for VL treatment, but the challenge is in ensuring that they are followed consistently across all facilities. There’s a lack of monitoring and support to make sure that every clinic is providing care according to these standards."*

Health service providers also expressed concerns about the overall health system structure, which they felt was not well equipped to handle the complexities of VL service delivery. A doctor shared, *"The health system here is very fragmented. There’s no clear chain of command, and sometimes it’s unclear who is responsible for what. This makes it difficult to implement guidelines effectively."*

### Financing: External and internal funding

The financing of VL services in Somalia is heavily dependent on external funding from international organizations and donors. This reliance on external sources of funding is both a strength and a weakness for VL service delivery.

An executive manager highlighted the importance of external funding: *"Our VL programs are almost entirely funded by international partners. Without their support, we wouldn’t be able to offer these services at all. The medicines, training, and even the salaries for some of our staff are covered by external funds."*

However, according to the participants, this dependency raises concerns about the sustainability of VL services. A health facility manager expressed this concern, saying, *"What worries us is what happens if the funding stops. There is no local funding to replace it, and that would mean the end of VL services here. It’s a constant source of anxiety."*

### INNER SETTING

#### Structural characteristics

The structural characteristics within the health facilities delivering visceral leishmaniasis (VL) services in Somalia, including physical infrastructure, information technology infrastructure, and work infrastructure, were identified as significant factors influencing service delivery. These structural components serve as both barriers and enablers, depending on their adequacy and functionality.

### Physical Infrastructure

The state of physical infrastructure was often expressed as a barrier to effective VL service delivery. Many health service providers and program managers expressed concerns about the inadequacy of the facilities. A health facility manager remarked, *"Most of our facilities are in poor condition. We lack basic amenities such as consistent electricity, clean water, and adequate space for patient care. This makes it very challenging to provide quality services, especially for a disease as complex as VL."*

Patients also expressed concerns about the physical conditions of the facilities. One patient shared, *"The clinic was overcrowded, and there were not enough beds. Some patients had to wait outside for their turn. It’s hard to feel confident in the treatment when the facility itself seems so rundown."*

### Information Technology Infrastructure

The lack of a robust information technology (IT) infrastructure was another barrier identified by participants. Health service providers emphasized the absence of electronic health records and communication systems that could streamline patient management and coordination across different levels of care. A qualified nurse stated, *"We still rely on paper records, and there is no system for tracking patients across different facilities. This makes it difficult to follow up on cases or share information between hospitals."*

### Work Infrastructure

Work infrastructure, including the availability of necessary tools, equipment, and materials, was also highlighted as a critical factor. Hospital managers pointed out that the lack of essential diagnostic tools, such as the Direct Agglutination Test (DAT) and microscopic examination capabilities, severely limits the ability to accurately diagnose VL. A hospital direcotr noted, *"Our labs are not equipped with the necessary tools to perform advanced diagnostic tests. We rely heavily on rapid tests, which are not always reliable. This compromises our ability to diagnose and treat VL effectively."*

### Communication

Effective communication within the health system was identified as a key enabler of VL service delivery, particularly in terms of integration with other services. Several participants highlighted the importance of communication and coordination between VL services and other health programs, such as malaria control, as a way to optimize resource use and improve patient outcomes.

An executive manager described the integration of VL and malaria services as a positive development: *"We’ve started integrating our VL services with malaria programs, and this has been very effective. The same teams that go out to diagnose and treat malaria can now also screen for VL. This has improved our coverage and allowed us to reach more patients."*

Health service providers also noted the benefits of improved communication within the health system. A doctor shared, *"The integration with malaria services has made it easier for us to identify VL cases early on. Patients who come in with fever are now routinely screened for both malaria and VL, which means we can start treatment sooner if needed."*

Patients also recognized the importance of these communication efforts. A patient mentioned, *"When I went to the clinic for malaria suspect, they also tested me for VL. I didn’t even know I had it until then. I’m grateful that they were able to catch it early."*

### Tension for Change

There was a palpable tension for change within the health facilities, with many participants expressing a desire for improvements in VL service delivery. This tension was driven by the recognition of existing gaps and the belief that better outcomes could be achieved with the right support and resources. A doctor noted, *"We know that the current system is not perfect, and there is a strong desire among the staff to make changes that will improve patient care. The challenge is finding the resources and support to make those changes happen."*

### Compatibility and Relative Priority

Participants highlighted the compatibility of VL services with other health programs and the relative priority given to VL within the broader health system. Program managers noted that while VL was recognized as an important health issue, it often competed with other priorities for attention and resources. An executive manager explained, *"VL is a serious disease, but it doesn’t always get the attention it deserves because there are so many other health issues that we’re dealing with. It’s a constant struggle to keep it on the agenda."*

Health service providers also expressed concerns about the relative priority given to VL. A doctor remarked, *"We’re often told that VL is important, but when it comes to allocating resources, it doesn’t always seem to be the top priority. This can be frustrating because we know how critical it is to address this disease."*

### Available Resources

The availability of resources, including funding, physical space, materials, and equipment, was identified as a crucial factor in the delivery of VL services. Participants consistently highlighted the challenges posed by limited resources, which often constrained their ability to provide effective care.

## Funding

The reliance on external funding for VL services was seen as both a strength and a weakness. Program managers expressed gratitude for the support provided by international partners but also voiced concerns about the sustainability of these funds. A Health manager explained, *"Our entire VL program is funded by external donors. While we’re grateful for their support, there’s always a concern about what will happen if the funding dries up. We don’t have a local funding base to fall back on."*

### Physical Space

The availability of physical space within health facilities was another challenge identified by participants. Many facilities were overcrowded, with limited space for patient care, diagnostic testing, and treatment. A qualified nurse noted, *"We simply don’t have enough space to accommodate all the patients who need VL treatment. Sometimes we have to turn people away or ask them to come back later, which is not ideal."*

### Materials and Equipment

The lack of necessary materials and equipment was a recurring theme among participants. Health service providers frequently mentioned the scarcity of diagnostic tools, medicines, and other essential supplies. A hospital director stated, *"We often run out of rapid tests we need for testing, and there are times when we don’t have the medicines we need to treat patients. This makes it very difficult to provide the level of care that our patients deserve."*

### Access to Knowledge and Information

Program managers and health service providers highlighted that continuous training and education regarding visceral leishmaniasis are not consistently available for healthcare providers. They also emphasized the importance of regular training to keep up with the latest developments in VL diagnosis and treatment. A health manager noted, *"Continuous education and training are crucial, but unfortunately, we don’t always have access to these resources. This gap in knowledge can affect the quality of care we provide"*.

Health service providers echoed this concern, with a doctor stating, *"We need ongoing training to stay updated on the best practices for diagnosing and treating VL. However, these opportunities are infrequent, which leaves us feeling unprepared at times."* The lack of continuous training and education was seen as a significant barrier to providing high-quality VL services, as it limits the ability of healthcare workers to stay informed about the latest advancements in treatment protocols and diagnostic techniques.

### INDIVIDUAL CHARACTERISTICS

#### Roles of high level leaders (Decision makers)

High-level leaders and decision-makers within the health system in terms of their commitment, vision, and leadership were expressed as a significant enabler on the implementation and sustainability of VL services. An executive manager highlighted the importance of strong leadership, stating, *"The commitment from our leadership team is vital. They have the influence to communicate and inform the partners to allocate resources, mobilize support, and ensure that VL services remain a priority. Without their backing, it would be difficult to sustain our efforts."*

However, the absence of such leadership can also serve as a barrier. A health manager noted, *"When leadership is not fully engaged or prioritizes other health issues, it becomes a challenge to maintain momentum for VL initiatives. We need more consistent advocacy from the top to keep VL on the agenda."*

### Roles of Opinion Leaders

Opinion leaders, who often serve as trusted voices within communities and the health system, were also seen as a key enabler who palys a pivotal role in influencing attitudes and behaviors related to VL service delivery. These individuals can act as champions for VL initiatives, advocating for their importance and helping to foster a supportive environment for implementation. A health facility manager mentioned, *"Opinion leaders in our community have been instrumental in raising awareness about VL. Their support helps us gain the trust of the community and encourages more people to seek care."*

#### 4.2.4 PROCESS

##### Planning

Particpants highlighted that effective planning ensures that resources are allocated appropriately, timelines are established, and goals are clearly defined. A program manager emphasized the importance of planning, stating, *"We need detailed plans to ensure that all aspects of VL service delivery are covered. Without proper planning, we risk delays and inefficiencies in implementation."*

A health manager noted, *"Having a clear work plan helps us stay organized and focused on our objectives. However, sometimes our plans are not detailed enough, leading to confusion and inconsistencies in service delivery."*

### Engaging

A program manager highlighted the role of engagement, stating, *"Engaging stakeholders helps us understand their perspectives and incorporate their feedback into our services. However, we often face challenges in effectively involving all relevant parties."*

Health service providers also shared their experiences, with one noting, *"Engaging with patients and community members helps build trust and ensures that our services meet their needs. But we sometimes struggle to engage all stakeholders effectively."*

### Reflecting and Evaluating

Reflecting and evaluating to assess the effectiveness of VL services, identifying areas for improvement, and making necessary adjustments based on feedback and outcomes were expressed as an important enabling factor. A health manager emphasized the importance of reflection and evaluation, stating, *"Regular evaluation helps us understand what is working and what needs improvement. Without it, we cannot effectively adapt our services to address emerging challenges."* Health service providers added, *"Reflecting on our practices and evaluating our outcomes allows us to make necessary changes and improve our service delivery. However, we often lack the resources and time to conduct thorough evaluations."*

### RECOMMENDED IMPLEMENTATION STRATEGIES TO IMPROVE VISCERAL LEISHMANIASIS SERVICE DELIVERY USING CFIR-ERIC MATCHING TOOL

By utilizing the CFIR-ERIC Matching tool, As provided in table 2, the study was able to translate the complex landscape of CFIR generated barriers into recommended strategies specifically tailored to the context of visceral leishmaniasis service delivery in Somalia. For example, CFIR barriers like complexity of VL services, particularly in diagnostic processes where healthcare providers struggled with limited tools and trainings were matched with strategies such as developing a formal implementation blueprint and promoting ongoing training to build local capacity.

**Table 2.** Implementation strategies matched using CFIR-ERIC Matching tool.

The systematic approach provided by the CFIR-ERIC Matching tool ensured that the strategies developed were not only theoretically sound but also practically feasible within the specific context of Somalia. This tailored approach is crucial for addressing the unique challenges faced in VL service delivery, enabling a more effective and sustainable implementation of services that can adapt to both the immediate and long-term needs of the population. The study’s use of the CFIR-ERIC tool highlights the importance of integrating evidence-based implementation strategies with a deep understanding of local barriers to improve health outcomes in complex settings like Somalia **(Table 2).**

## DISCUSSION

The study examined the implementation of visceral leishmaniasis services in somalia, identifying key barriers and enablers using the determinant framework CFIR. The results showed in the study provide a detailed understanding of the complexities sorround with delvering effective VL services in a resource limited and conflict effected setting like somalia.

Adaptability issues, such as rigid treatment protocols failing to account for local realities like drug shortages and patient mobility, complexity in diagnostics and treatment, exacerbated by inadequate training and tools were a main challenge. Other countries in east Africa had the same adaptation and complexity problem in terms of adapting the services to the local context, supply chain of diagnostic processes and access to treatment [27]. In our study, the cost associated with implementing VL services was also identified as a significant barrier. This includes both the direct costs of procuring drugs, diagnostic tools, and other essential supplies, as well as the indirect costs associated with training healthcare providers, maintaining infrastructure, and conducting surveillance. The reliance on international partners for funding and support was highlighted as a vulnerability. A similar study on barriers to treatment for visceral leishmaniasis in hyperendemic areas including India, Brazil, Nepal, Bangladesh and Sudan revealed similar instances regarding the cost of treatment for visceral leishmaniasis [44].

### 5.2 OUTER SETTING

The study found that critical incidents, such as emergencies and outbreaks, can significantly disrupt VL service delivery. Program managers and healthcare providers reported that during periods of conflict or natural disasters, resources are often diverted away from VL services to address more immediate concerns. Local attitudes and sociocultural factors also play a significant role in the implementation of VL services. An earlier review found that challenges related with sociocultural beliefs, poor health system and political instability in eastern African regions impede the implementation of effective visceral leishmaniasis control [45]. Another study noted that conflicts contribute to increase in cases of visceral leishmaniasis through population displacement and health system deterioration [46]. Our study also found that there are widespread misconceptions about VL among the population, leading to delays in seeking treatment. Patients often rely on traditional healers or self-medication before seeking medical care, which can result in advanced disease and poorer outcomes. Program managers noted that these attitudes are influenced by a lack of awareness about VL and the availability of treatment. A study in Sudan revealed that misconceptions about VL were the main driving factors for delay in earlier seeking treatment [47]. In this study, we found that collaboration and partnership with international organizations has provided essential funding, technical support, and resources for VL services in Somalia. Program managers emphasized the importance of building local partnerships and referral networks to enhance the sustainability of VL services. We also found that the regulatory environment in Somalia is not fully supportive of VL service delivery. Program managers reported that there are gaps in the existing policies and laws, which do not adequately address the specific needs of VL control. Additionally, the overall health system structure, characterized by limited coordination and fragmentation, hampers the effective implementation of VL services. The study revealed that financing is a significant barrier to VL service delivery, with funding often coming from external entities. This dependence on external funding creates uncertainty and limits the ability to plan and implement long-term VL control strategies. Program managers also highlighted the impact of external pressure, including social pressure from mass media campaigns and advocacy groups, which can influence the prioritization of VL services. An earlier study in east Africa confirms that challenges related with coordination, stable funding, regulatory requirements, and overall health system structure hamper effective implementation of visceral leishmaniasis service delivery [27].

### 5.3 INNER SETTING

The study found that the physical infrastructure, information technology infrastructure, and work infrastructure in Somalia are inadequate to support effective VL service delivery. Healthcare facilities often lack the necessary equipment, space, and technological support to provide comprehensive VL care. Communication within the health system was also identified as a barrier. The study found that there are significant gaps in communication between different levels of the health system, which leads to inefficiencies and delays in service delivery. Program managers reported that the lack of clear communication channels and protocols often results in confusion and misalignment of priorities. Similar communication and infrastructure challenges were also found in other regions including east Africa and Asia [48]. Relative priority and incentive systems were also identified as barriers in the inner setting domain. The study found that VL services are often not prioritized within the health system, which leads to limited resources and support for these services. Program managers reported that other health issues, such as malaria and tuberculosis, often take precedence over VL, which affects the allocation of resources and attention. The study also found that there is a significant gap in access to knowledge and information within the health system. Healthcare providers and program managers reported that they often lack access to the latest guidelines, research, and best practices for VL service delivery.

### 5.4 INDIVIDUAL CHARACTERISTICS

The study highlighted a complex interplay among various stakeholders and highlighted critical roles in shaping the effectiveness of VL services. For instance, the high-level leaders and decision-makers were identified as crucial enablers of VL service implementation. Their commitment, vision, and leadership were seen as essential for sustaining VL initiatives and mobilizing resources. Conversely, the absence of engaged leadership often acted as a barrier, as noted by a health manager who emphasized the difficulties faced when leadership shifted focus to other health issues. Opinion leaders, including respected figures within communities and the health system, were also pivotal in VL service delivery. These individuals served as champions for VL initiatives, influencing attitudes and behaviors related to VL care. Their support was instrumental in raising awareness and fostering a supportive environment for the implementation of VL services. A health facility manager acknowledged the critical role of opinion leaders in gaining community trust and encouraging care-seeking behavior. Other individual roles were also identified as significant enablers in effective implementation of visceral leishmaniasis service delivery.

### 5.5 PROCESS

Regarding the implementation process of delivering visceral leishmaniasis service delivery, the study revealed that assessing the needs and context was a critical process. Planning was identified as a vital process for ensuring the effective delivery of VL services. A program manager stressed the importance of having comprehensive plans to cover all aspects of VL service delivery. Engagement with stakeholders was highlighted as crucial for incorporating feedback and understanding different perspectives. A program manager observed that engaging stakeholders helped in refining services based on their input. However, challenges in involving all relevant parties effectively were noted. Reflecting and evaluating VL services were essential for assessing their effectiveness and making necessary adjustments. A health manager emphasized that regular evaluation was vital for understanding what worked well and what needed improvement. Without systematic reflection and evaluation, it was challenging to adapt services to emerging challenges.

### 5.6 CONCLUSION

The study provides a comprehensive analysis on the current status, availability, accessibility, barriers and enablers to effective implementation of visceral leishmaniasis service delivery in somalia. The study also highlights recommended strategies to develop an effective implementation strategy to improve the VL service delivery in the country. By addressing identified gaps, its possible to improve and implement the recommended strategies to achieve elimination of visceral leishmaniasis as a public health problem in Somalia. This requires a multi-faceted approach, involving strong leadership, tailored interventions, continuous training, and effective engagement of all stakeholders.

## Data Availability

All data produced in the present study are available upon reasonable request to the authors

## Acknowledgements

The authors wish to express their sincere gratitude to all stakeholders who contributed to this study, including executive directors, program managers, government entities, healthcare workers, and community members in the study areas. Special thanks to the staff at the participating healthcare facilities for their cooperation and support during data collection.

## Author Contributions

**Abdirahman Moallim Ibrahim:** Conceptualization, Methodology, Investigation, Data Curation, Writing - Original Draft. **Lul Mohamud Mohamed:** Formal Analysis, Validation, Writing - Review & Editing. **Anoop Khanna:** Supervision, Methodology, Review & Editing. **Mohamed Mohamud Fuje:** Investigation, Resources, Data Curation. **Liban Hassan Jimale:** Validation, Writing - Review & Editing. **Mohamed Abdirahman Omer:** Validation, Writing - Review & Editing. **Rimaz Mohamed Dewelbait:** Formal Analysis, Visualization, Writing - Review & Editing.

## Disclosure Statement

The authors report no conflicts of interest. The authors alone are responsible for the content and writing of this paper.

## Consent for Publication

Not applicable.

## Ethics Approval and Consent to Participate

This study was conducted in accordance with the ethical principles of the Helsinki Declaration. Ethical approval was obtained from the Somali National Institute of Health Institutional Review Board (Reference: **NIH/IRB/11/DEC/2023**). Written informed consent was obtained from all study participants, and confidentiality of all information was maintained throughout the research process.

## Funding

This research received no specific grant from any funding agency in the public, commercial, or not-for-profit sectors.

## Patient and Public Involvement

Patients and the public were not directly involved in the design or conduct of this study due to its exploratory qualitative nature. However, study findings will be disseminated to relevant patient advocacy groups and community stakeholders through appropriate channels.

## Data Availability

The datasets used and analyzed during the current study are available from the corresponding author upon reasonable request.

## Notes

### Competing Interest Statement

The authors have declared no competing interest.

### Author Declarations

Ethical approval was obtained from the Somali National Institute of Health Institutional Review Board (Reference: NIH/IRB/11/DEC/2023).

## References

1. WHO. Leishmaniasis [Internet]. 2023 [cited 2024 Aug 12]. Available from: https://www.who.int/news-room/fact-sheets/detail/leishmaniasis

2. Leishmaniasis - ScienceDirect [Internet]. [cited 2024 Aug 12]. Available from: https://www.sciencedirect.com/science/article/abs/pii/B978008055232360933X

3. CDC. About Leishmaniasis [Internet]. Leishmaniasis. 2024 [cited 2024 Aug 12]. Available from: https://www.cdc.gov/leishmaniasis/about/index.html

4. Symptoms, transmission, and current treatments for visceral leishmaniasis | DNDi [Internet]. 2020 [cited 2024 Aug 12]. Available from: https://dndi.org/diseases/visceral-leishmaniasis/facts/

5. Scarpini S, Dondi A, Totaro C, Biagi C, Melchionda F, Zama D, et al. Visceral Leishmaniasis: Epidemiology, Diagnosis, and Treatment Regimens in Different Geographical Areas with a Focus on Pediatrics. Microorganisms. 2022 Sep 21;10(10):1887.

6. Visceral Leishmaniasis - PAHO/WHO | Pan American Health Organization [Internet]. [cited 2024 Aug 12]. Available from: https://www.paho.org/en/topics/leishmaniasis/visceral-leishmaniasis

7. Wamai RG, Kahn J, McGloin J, Ziaggi G. Visceral leishmaniasis: a global overview. J Glob Health Sci [Internet]. 2020 May 14 [cited 2024 Aug 12];2(1). Available from: 10.35500/jghs.2020.2.e3

8. Ready PD. Full article: Epidemiology of visceral leishmaniasis [Internet]. [cited 2024 Aug 12]. Available from: https://www.tandfonline.com/doi/full/10.2147/CLEP.S44267

9. Sunyoto T, Potet J, Boelaert M. Visceral leishmaniasis in Somalia: A review of epidemiology and access to care. PLoS Negl Trop Dis. 2017 Mar;11(3):e0005231.

10. UNFPA Somalia. Somali Health and Demographic Survery [Internet]. 2020. Available from: https://somalia.unfpa.org/sites/default/files/pub-pdf/FINAL%20SHDS%20Report%202020_V7_0.pdf

11. Gele A. Challenges Facing the Health System in Somalia and Implications for Achieving the SDGs. Eur J Public Health. 2020 Sep 1;30(Supplement_5):ckaa165.1147.

12. Pigott DM, Bhatt S, Golding N, Duda KA, Battle KE, Brady OJ, et al. Global distribution maps of the leishmaniases. Tollman S, editor. eLife. 2014 Jun 27;3:e02851.

13. Visceral leishmaniasis in eastern Africa – current status - PMC [Internet]. [cited 2024 Aug 12]. Available from: https://www.ncbi.nlm.nih.gov/pmc/articles/PMC2664918/

14. Jones CM, Welburn SC. Leishmaniasis Beyond East Africa. Front Vet Sci [Internet]. 2021 Feb 26 [cited 2024 Aug 12];8. Available from: https://www.frontiersin.org/journals/veterinary-science/articles/10.3389/fvets.2021.618766/full

15. Ahmed M, Abdulslam Abdullah A, Bello I, Hamad S, Bashir A. Prevalence of human leishmaniasis in Sudan: A systematic review and meta-analysis. World J Methodol. 2022 Jul 20;12(4):305–18.

16. Naylor-Leyland G, Collin SM, Gatluak F, Boer M den, Alves F, Mullahzada AW, et al. The increasing incidence of visceral leishmaniasis relapse in South Sudan: A retrospective analysis of field patient data from 2001–2018. PLoS Negl Trop Dis. 2022 Aug 18;16(8):e0010696.

17. Leta S, Dao THT, Mesele F, Alemayehu G. Visceral leishmaniasis in Ethiopia: an evolving disease. PLoS Negl Trop Dis. 2014 Sep;8(9):e3131.

18. Grifferty G, Shirley H, O’Brien K, Hirsch JL, Orriols AM, Amechi KL, et al. The leishmaniases in Kenya: A scoping review. PLoS Negl Trop Dis. 2023 Jun 1;17(6):e0011358.

19. Olobo-Okao J, Sagaki P. Leishmaniasis in Uganda: Historical account and a review of the literature. Pan Afr Med J. 2014 May 4;18:16.

20. Bekele F, Belay T, Zeynudin A, Hailu A. Visceral leishmaniasis in selected communities of Hamar and Banna-Tsamai districts in Lower Omo Valley, South West Ethiopia: Sero- epidemological and Leishmanin Skin Test Surveys. PloS One. 2018;13(5):e0197430.

21. Ketema H, Weldegebreal F, Gemechu A, Gobena T. Seroprevalence of visceral leishmaniasis and its associated factors among asymptomatic pastoral community of Dire District, Borena zone, Oromia Region, Ethiopia. Front Public Health [Internet]. 2022 Nov 21 [cited 2024 Aug 12];10. Available from: https://www.frontiersin.org/journals/public-health/articles/10.3389/fpubh.2022.917536/full

22. Alebie G, Worku A, Yohannes S, Urga B, Hailu A, Tadesse D. Epidemiology of visceral leishmaniasis in Shebelle Zone of Somali Region, eastern Ethiopia | Parasites & Vectors | Full Text [Internet]. [cited 2024 Aug 12]. Available from: https://parasitesandvectors.biomedcentral.com/articles/10.1186/s13071-019-3452-5

23. Mewara A, Gudisa R, Padhi BK, Kumar P, Sah R, Rodriguez-Morales AJ. Visceral leishmaniasis outbreak in Kenya—a setback to the elimination efforts. New Microbes New Infect. 2022 Nov 29;49–50:101060.

24. Marlet MVL, Sang DK, Ritmeijer K, Muga RO, Onsongo J, Davidson RN. Emergence or re-emergence of visceral leishmaniasis in areas of Somalia, north-eastern Kenya, and south- eastern Ethiopia in 2000-01. Trans R Soc Trop Med Hyg. 2003 Oct;97(5):515–8.

25. Al-Salem W, Herricks JR, Hotez PJ. A review of visceral leishmaniasis during the conflict in South Sudan and the consequences for East African countries. Parasit Vectors. 2016 Aug 22;9(1):460.

26. Boodman C, Griensven J van, Gupta N, Diro E, Ritmeijer K. Anticipating visceral leishmaniasis epidemics due to the conflict in Northern Ethiopia - PMC [Internet]. 2023 [cited 2024 Aug 12]. Available from: https://www.ncbi.nlm.nih.gov/pmc/articles/PMC10035745/

27. Sunyoto T, Potet J, den Boer M, Ritmeijer K, Postigo JAR, Ravinetto R, et al. Exploring global and country-level barriers to an effective supply of leishmaniasis medicines and diagnostics in eastern Africa: a qualitative study. BMJ Open. 2019 May 30;9(5):e029141.

28. Alvar J, Beca-Martínez MT, Argaw D, Jain S, Aagaard-Hansen J. Social determinants of visceral leishmaniasis elimination in Eastern Africa. BMJ Glob Health. 2023 Jun 1;8(6):e012638.

29. Elnaiem DEA. Ecology and control of the sand fly vectors of Leishmania donovani in East Africa, with special emphasis on Phlebotomus orientalis. J Vector Ecol. 2011 Mar 2;36:S23–31.

30. TropicalMed | Free Full-Text | Infection of Leishmania donovani in Phlebotomus orientalis Sand Flies at Different Microhabitats of a Kala-Azar Endemic Village in Eastern Sudan [Internet]. [cited 2024 Aug 12]. Available from: https://www.mdpi.com/2414-6366/9/2/40

31. Control of visceral leishmaniasis in Somalia: achievements in a challenging scenario, 2013– 2015. Releve Epidemiol Hebd. 2017 Sep 22;92(35):566–72.

32. Guha-Sapir D, Ratnayake R. Consequences of Ongoing Civil Conflict in Somalia: Evidence for Public Health Responses. PLOS Med. 2009 Aug 11;6(8):e1000108.

33. Raguenaud ME, Jansson A, Vanlerberghe V, Deborggraeve S, Dujardin JC, Orfanos G, et al. Epidemiology and clinical features of patients with visceral leishmaniasis treated by an MSF clinic in Bakool region, Somalia, 2004-2006. PLoS Negl Trop Dis. 2007 Oct 31;1(1):e85.

34. Marlet MVL, Wuillaume F, Jacquet D, Quispe KW, Dujardin JC, Boelaert M. A neglected disease of humans: a new focus of visceral leishmaniasis in Bakool, Somalia. Trans R Soc Trop Med Hyg. 2003 Dec;97(6):667–71.

35. WHO. Service availability and readiness assessment (SARA) [Internet]. 2015 [cited 2024 Aug 14]. Available from: https://www.who.int/data/data-collection-tools/service-availability-and-readiness-assessment-(sara)

36. The Consolidated Framework for Implementation Research – Technical Assistance for users of the CFIR framework [Internet]. [cited 2024 Aug 14]. Available from: https://cfirguide.org/

37. Yakovchenko V, Chinman MJ, Lamorte C, Powell BJ, Waltz TJ, Merante M, et al. Refining Expert Recommendations for Implementing Change (ERIC) strategy surveys using cognitive interviews with frontline providers. Implement Sci Commun [Internet]. 2023 [cited 2024 Aug 14];4. Available from: https://www.ncbi.nlm.nih.gov/pmc/articles/PMC10122282/

38. Damschroder LJ, Reardon CM, Widerquist MAO, Lowery J. The updated Consolidated Framework for Implementation Research based on user feedback | Implementation Science | Full Text [Internet]. 2022 [cited 2024 Aug 14]. Available from: https://implementationscience.biomedcentral.com/articles/10.1186/s13012-022-01245-0

39. Damschroder LJ, Aron DC, Keith RE, Kirsh SR, Alexander JA, Julie C Lowery. Fostering implementation of health services research findings into practice: a consolidated framework for advancing implementation science | Implementation Science | Full Text [Internet]. 2009 [cited 2024 Aug 14]. Available from: https://implementationscience.biomedcentral.com/articles/10.1186/1748-5908-4-50

40. Damschroder LJ, Reardon CM, Opra Widerquist MA, Lowery J. Conceptualizing outcomes for use with the Consolidated Framework for Implementation Research (CFIR): the CFIR Outcomes Addendum. Implement Sci. 2022 Dec;17(1):1–10.

41. CFIR-ERIC Implementation Strategy Matching Tool | NCCMT [Internet]. [cited 2024 Aug 14]. Available from: https://www.nccmt.ca/organizational-change/results/77

42. Powell BJ, Waltz TJ, Chinman MJ, Damschroder LJ, Smith JL, Matthieu MM, et al. A refined compilation of implementation strategies: results from the Expert Recommendations for Implementing Change (ERIC) project. Implement Sci. 2015 Dec;10(1):1–14.

43. Waltz TJ, Powell BJ, Matthieu MM, Damschroder LJ, Chinman MJ, Smith JL, et al. Use of concept mapping to characterize relationships among implementation strategies and assess their feasibility and importance: results from the Expert Recommendations for Implementing Change (ERIC) study. Implement Sci. 2015 Dec;10(1):1–8.

44. Thornton SJ, Wasan KM, Piecuch A, Lynd LLD, Wasan EK. Barriers to treatment for visceral leishmaniasis in hyperendemic areas: India, Bangladesh, Nepal, Brazil and Sudan. Drug Dev Ind Pharm [Internet]. 2010 Nov 1 [cited 2024 Aug 15]; Available from: https://www.tandfonline.com/doi/abs/10.3109/03639041003796648

45. Makau-Barasa LK, Ochol D, Yotebieng KA, Adera CB, de Souza DK. Moving from control to elimination of Visceral Leishmaniasis in East Africa. Front Trop Dis. 2022 Aug 19;3:965609.

46. Berry I, Berrang-Ford L. Leishmaniasis, conflict, and political terror: A spatio-temporal analysis. Soc Sci Med. 2016 Oct 1;167:140–9.

47. Sunyoto T, Adam GK, Atia AM, Hamid Y, Babiker RA, Abdelrahman N, et al. “Kala-Azar is a Dishonest Disease”: Community Perspectives on Access Barriers to Visceral Leishmaniasis (Kala-Azar) Diagnosis and Care in Southern Gadarif, Sudan. Am J Trop Med Hyg. 2018 Apr;98(4):1091–101.

48. Hailu T, Yimer M, Mulu W, Abera B. Challenges in visceral leishmaniasis control and elimination in the developing countries: A review. J Vector Borne Dis. 2016 Sep;53(3):193– 8.

