## Supplementary material for "Improving Visceral Leishmaniasis Service Delivery In Somalia: An Exploratory Qualitative Study Informed By CFIR-ERIC Matching Tool": Suplementary files: COREQ_Checklist.docx

**Supporting Information**

Consolidated criteria for reporting qualitative studies (COREQ): 32-item checklist.

| **No** | **Item** | **Guide Questions/Description** | **Page Number** |
| --- | --- | --- | --- |
| **Domain 1: Research team and reflexivity** | | | |
| **Personal Characteristics** | |  |  |
| 1. | Interviewer/facilitator | Abdirahman Moallim Ibrahim | 5 |
| 2. | Credentials | MBChB, MPH(IS) | 5 |
| 3. | Occupation | Research officer, Jazeera University | 5 |
| 4. | Gender | The interviewer was male | NA |
| 5. | Experience and training | AMI, LMM, AK, and RMD have experience and trained for conducting in-depth interviews and writing for qualitative research studies. | 5 |
| **Relationship with participants** | | | |
| 6. | Relationship established | None. | 5 |
| 7. | Participant knowledge of the interviewer | Few participants were known to the AMI | 5 |
| 8. | Interviewer characteristics | The interviewer is a public health and medical researcher and is particularly interested in qualitative and mixed methods | 5 |
| **Domain 2: Study design** | | | |
| **Theoretical framework** | |  |  |
| 9. | Methodological orientation and Theory | Qualitative methods and thematic analysis | 6 |
| 10. | Sampling | Purposive sampling | 5 |
| 11. | Method of approach | We approach stakeholders through LinkedIn, email contacts and phone contact. After invitation and agreement, we conducted all the interviews through face to face | 5 |
| 12. | Sample size | 57 for IID and 16 for FGD | 5 |
| 13. | Non-participation | None. We purposively selected participants for interviews. And all participants agreed to give an interview after we approached them. | 5 |
| **Setting** | | | |
| 14. | Setting of data collection | In person | 7 |
| 15. | Presence of non-participants | No | 7 |
| **Data collection** | | | |
| 17. | Interview guide: An interview guide from CFIR model was drafted and revised. | | 7 |
| 18. | Repeat interviews | None | 7 |
| 19. | Audio/visual recording | All interviews were audio-recorded and there were no field visits conducted | 7 |
| 20. | Field notes | Yes | 7 |
| 21. | Duration | 45 mins-60 minutes | 7 |
| 22. | Data saturation | Yes | 7 |
| 23. | Transcripts returned | No | 7 |
| **Domain 3: Analysis and findings** | | | |
| **Data analysis** | | | |
| 24. | Number of data coders | 3 | 8 |
| 25. | Description of the coding tree | CFIR Domains was used | 8 |
| 26. | Derivation of themes | The theme was derived from CFIR based on thematic analysis approach | 8 |
| 27. | Software | EXCEL Software | 8 |
| 28. | Participant checking | Sought informal feedback from some of the participants to check the validity of the results. | 8 |
| **Reporting** | | | |
| 29. | Quotations presented | Yes | 9-20 |
| 30. | Data and findings consistent | Several relevant quotations used to illustrate findings | 9-20 |
| 31. | Clarity of major themes | Yes | 9-20 |
| 32. | Clarity of minor themes | Yes | 9-20 |
