## Supplementary material for "Improving Visceral Leishmaniasis Service Delivery In Somalia: An Exploratory Qualitative Study Informed By CFIR-ERIC Matching Tool": Suplementary files: Interview guide for IID and FGD.docx

**VISCERAL LEISHMANIASIS CHECKLIST AND INTERVIEW GUIDE**

**Name of facility: _______________________________________**

**Location of facility: _____________________________________**

**Region/province: ______________________________________**

**Type of facility: ________________________________________**

**Dates of Visits (in dd/mm/yyyy): _________________________**

**Urban/rural: __________________________________________**

**Visceral leishmaniasis**

**PART I- Service Statistics (This needs to be filled in by the concerned facility)**

|  | **Employed** | **Unemployed** |
| --- | --- | --- |
| **Staffing** |  |  |
| - General medical doctors trained for VL diagnosis and treatment |  |  |
| - Specialist medical doctors |  |  |
| - Nursing professionals |  |  |
| - Laboratory technicians |  |  |
| - Pharmacists |  |  |
| - Community Health Workers |  |  |
| **Inpatient and observation beds** |  |  |
| 1) Excluding any delivery beds, how many overnight/inpatient beds in total does this facility have, both for adults and children? |  |  |
| 2) Of the overnight/inpatient beds in this facility, how many are dedicated VL beds? |  |  |
| **Infrastructure** | **Yes** | **No** |
| - Does this facility have a functioning computer? |  |  |
| - Is there access to email or internet within the facility today? |  |  |
| - Does this facility have a functional ambulance or other vehicle for emergency transportation for clients that is stationed at this facility or operates from this facility? |  |  |
| - Is fuel for the ambulance or other emergency vehicle available today? |  |  |
| - Does your facility have electricity from any source (e.g., electricity grid, generator, solar, or other)? |  |  |
| - Is there a designated area for waste disposal? |  |  |
| - Are there microscopes available for VL diagnosis? |  |  |
| - Are there centrifuges available for VL diagnosis? |  |  |
| - Are there refrigerators available to store VL diagnostic reagents? |  |  |
| - Are there other essential equipment for VL diagnosis and treatment available, such as RDTs, personal protective equipment, and syringes? |  |  |
| - Does this facility have any guidelines on standard precautions for infection prevention? |  |  |
| **Basic amenities** |  |  |
| - On average, how many hours per day is this facility open? | 1) **4 hours to 5 hours** |  |
|  | 2) **6 hours to 8 hours** |  |
|  | 3) **9 hours to 12 hours** |  |
|  | 4) **13 hours to 24 hours** |  |
|  | 5) **More than 24 hours** |  |
| **Medicines and commodities** | **Yes** | **No** |
| - Are essential medicines for VL treatment available, such as antiparasitic drugs? |  |  |
| - Are laboratory reagents for VL diagnosis available? |  |  |
| - Are personal protective equipment for healthcare providers available? |  |  |
| **Diagnostics** | **Yes** | **No** |
| - Are RDTs for VL diagnosis available? |  |  |
| - Are other diagnostic tests for VL available, such as PCR? |  |  |
| **Training** | **Yes** | **No** |
| - Are healthcare providers (medical doctors and laboratory technicians) trained in VL diagnosis and treatment for the last 6 months? |  |  |
| - Are healthcare providers up to date on the latest VL treatment guidelines? |  |  |
| - Are there opportunities for continuing medical education (CME) on VL for healthcare providers? |  |  |
| **Readiness** | **Yes** | **No** |
| - Are there standard operating procedures (SOPs) in place for VL diagnosis and treatment? |  |  |
| - Is there a referral mechanism for patients with complicated VL cases? |  |  |
| - Is there a data collection system in place to track VL cases? |  |  |
| **Accessibility** | **Yes** | **No** |
| - Are VL diagnosis and treatment services available within a reasonable distance for all patients? |  |  |
| - Is there adequate transportation available for patients to access VL diagnosis and treatment services? |  |  |
| - Are VL diagnosis and treatment services affordable for all patients? |  |  |
| - Is there a fee-for-service system for VL diagnosis and treatment? |  |  |
| - Are there any subsidies or exemptions available for VL diagnosis and treatment? |  |  |
| - Are healthcare providers culturally sensitive to the needs of VL patients? |  |  |
| - Are there language interpreters available to assist patients who do not speak the local language? |  |  |
| - Are there culturally appropriate communication materials available for patients? |  |  |
|  | **Yes** | **No** |
| - Are RDTs for VL diagnosis accurate? |  |  |
| - Is microscopy for VL diagnosis accurate? |  |  |
| - Are other diagnostic tests for VL accurate? |  |  |
| - Are essential medicines for VL treatment effective? |  |  |
| - Are the treatment regimens for VL effective? |  |  |
| - Is there a system in place to monitor treatment outcomes? |  |  |
| - Are patients satisfied with the quality of VL diagnosis and treatment services? |  |  |
| - Are patients satisfied with the communication from healthcare providers? |  |  |
| - Are patients satisfied with the overall experience of receiving VL diagnosis and treatment? |  |  |

**Part III.A – Provider Interaction**

**A. Provider Interaction – Service provider**

**I. Demographics**

1. Age: _________	Yrs.: _____________

2. Gender: __________

3. Current affiliation and designation: _______________________________

4. Can you describe the availability of VL diagnosis and treatment services at your facility?

______________________________________________________________________________________________________________________________________________________________________________________________________________________________________________________________________________________________________________________________________________________________________________________________________

5. What are the main challenges you face in providing VL diagnosis and treatment services at your facility?

________________________________________________________________________________________________________________________________________________________________________________________________________________________________________________________________________________________________________________________

6. What are the available resources and infrastructure for VL diagnosis and treatment at your facility?

________________________________________________________________________________________________________________________________________________________________________________________________________________________________________________________________________________________________________________________

7. What training and support have you received in managing VL cases?

________________________________________________________________________________________________________________________________________________________________________________________________________________________________________________________________________________________________________________________

8. What factors, from your perspective, hinder effective VL service delivery?

________________________________________________________________________________________________________________________________________________________________________________________________________________________________________________________________________________________________________________________________________________________

9. What factors, in your opinion, facilitate the improvement of VL service delivery?

________________________________________________________________________________________________________________________________________________________________________________________________________________________________________________________________________________________________________________________________________________________

10. What changes or interventions do you believe would significantly enhance VL service delivery?

________________________________________________________________________________________________________________________________________________________________________________________________________________________________________________________________________________________________________________________________________________________

11. What role do you think technology can play in improving VL service delivery?

________________________________________________________________________________________________________________________________________________________________________________________________________________________________________________________________________________________________________________________________________________________

12. What suggestions do you have for enhancing the collaboration between health providers, patients, and decision-makers?

________________________________________________________________________________________________________________________________________________________________________________________________________________________________________________________________________________________________________________________

13. How do you collaborate with other healthcare providers, such as community health workers, to provide VL services?

________________________________________________________________________________________________________________________________________________________________________________________________________________________________________________________________________________________________________________________

**PART III B. Decision makers & partners –MoH, MSF, SOS, UNICEF, WHO**

1. Can you provide an overview of the current state of VL service delivery in Somalia?

____________________________________________________________________________________________________________________________________________________________________________________________________________________________________________________________________________

2. What are the key challenges and opportunities in improving VL control at a national level?

________________________________________________________________________________________________________________________________________________________________________________________________________________________________________________________________________________________________

3. How are resources allocated for VL control and service delivery?

________________________________________________________________________________________________________________________________________________________________________________________________________________________________________________________________________________________________

4. What role does the Ministry of Health (MOH) play in overseeing and regulating VL control efforts?

___________________________________________________________________________________________________________________________________________________________________________________________________________________________________________________________________________________________________________________________

5. How do international NGOs collaborate with the MOH and other stakeholders to improve VL control?ss

____________________________________________________________________________________________________________________________________________________________________________________________________________________________________________________________________________________________________________________________

6. What are the potential barriers and facilitators to implementing effective VL control strategies?

________________________________________________________________________________________________________________________________________________________________________________________________________________________________________________________________________________________________

7. What are the top priorities for improving VL service delivery in Somalia?

________________________________________________________________________________________________________________________________________________________________________________________________________________________________________________________________________________________________

8. What mechanisms are in place to monitor and evaluate the effectiveness of VL control interventions?

________________________________________________________________________________________________________________________________________________________________________________________________________________________________________________________________________________________________

9. How can evidence-based recommendations be translated into effective VL control programs?

________________________________________________________________________________________________________________________________________________________________________________________________________________________________________________________________________________________________

**Part IV**

**Community Interaction –Patient interaction**

|  | **1** | **2** | **3** |
| --- | --- | --- | --- |
| **Age of the patient** |  |  |  |
| **Gender** | **M** 🞎 F 🞎 | **M** 🞎 F 🞎 | **M** 🞎 F 🞎 |
| **Resident of the patient** |  |  |  |
| 1. **Can you describe your experiences with accessing and receiving VL diagnosis and treatment services?** |  |  |  |
| 2. **What were the barriers you faced in accessing and receiving VL diagnosis and treatment services?** |  |  |  |
| 3. **What aspects of your experiences with VL diagnosis and treatment services could be improved?** |  |  |  |
| 4. **Can you describe the challenges you faced during your interactions with healthcare providers?** |  |  |  |
| 5. **What were the factors that influenced your decision to seek VL diagnosis and treatment?** |  |  |  |
| 6. **How did you learn about VL and its symptoms?** |  |  |  |
| 7. **What steps did you take to seek diagnosis and treatment?** |  |  |  |
| 8. **What were your experiences with transportation and logistics in accessing VL services?** |  |  |  |
| 9. **How did the cost of VL diagnosis and treatment affect your decision to seek care?** |  |  |  |
| 10. **What suggestions do you have for improving the patient experience with VL diagnosis and treatment services?** |  |  |  |
