## Supplementary material for "Improving Visceral Leishmaniasis Service Delivery In Somalia: An Exploratory Qualitative Study Informed By CFIR-ERIC Matching Tool": Suplementary files: Tables 1&2.docx

**Table 1 Status of visceral leishmaniasis centers**

|  | **Visceral leishmaniasis centers** | | | | |
| --- | --- | --- | --- | --- | --- |
|  | **SOS Mogadishu** | **De Martino Hospital** | **Benaadir Hospital** | **SOS Baidoa** | **Bay Regional Hospital** |
| **Staffing** | +++ | +++ | +++ | +++ | +++ |
| **Inpatient and observation beds** | +++ | ++ | +++ | +++ | +++ |
| **Infrastructure** | ++ | ++ | ++ | +++ | ++ |
| **Basic amenities** | +++ | +++ | +++ | +++ | +++ |
| **Medicines and commodities** | +++ | + | +++ | +++ | ++ |
| **Diagnostics** | +++ | + | ++ | +++ | + |
| **Information system** | +++ | + | ++ | +++ | + |

**Table 2 Implementation strategies matched using CFIR-ERIC Matching tool**

| **CFIR Barriers** | **ERIC Strategies** | **Ranks (%) **** | **Proposed context-specific strategies** |
| --- | --- | --- | --- |
| **Adaptability** | Promote adaptability | 73 | Develop flexible VL service delivery models that adapt to changing local conditions and patient needs, such as mobile clinics and outreach teams. |
| **Complexity** | Develop a formal implementation blueprint | 43 | Create detailed guidelines for VL service delivery and provide continuous training to health workers, ensuring they can adapt to complex cases. |
|  | Promote adaptability | 40 |  |
|  | Conduct ongoing training | 37 |  |
| **Cost of Implementing VL services** | Access new funding | 72 | Secure external funding from, domestic, NGOs and international donors to reduce the cost burden. |
|  | Alter incentive/allowance structures | 44 | Adjust healthcare provider incentives to improve service delivery. |
| **Critical incidents: emergencies and outbreaks** |  |  |  |
| **Sociocultural beliefs and values** |  |  |  |
| **Economic, environmental and political conditions** |  |  |  |
| **Partnerships, referral networks and Connections** | Build a coalition | 62 | Strengthen referral networks between health facilities, build coalitions with NGOs, and form academic partnerships for technical and research support. |
|  | Develop academic partnerships | 50 |  |
|  | Promote network weaving | 50 |  |
|  | Use advisory boards and workgroups | 35 |  |
|  | Visit other sites | 35 |  |
| **Policies, guidelines and health system structure** |  |  | Advocate for national policies and guidelines that prioritize VL service delivery and integrate it into the routine health system. |
| **Internal and external funding** |  |  | Strengthen partnerships with NTD donors and government entities to secure consistent funding for VL services. |
| **Competing priorities** |  |  | Elevate VL as a public health priority through advocacy and awareness campaigns to ensure it receives adequate resources and attention. |
| **Culture-centered equity and patient care** | Obtain and use patients/consumers and family feedback | 76 | Involve patients and their families in care decisions and use their feedback to improve services. Conduct community-driven needs assessments to tailor interventions. |
|  | Involve patients/consumers and family members | 71 |  |
|  | Conduct local needs assessment | 57 |  |
|  | Prepare patients/consumers to be active participants | 48 |  |
|  | Conduct local consensus discussions | 29 |  |
| **Structural Characteristics: physical, information technology and work infrastructure** | Assess for readiness and identify barriers and facilitators | 36 | Upgrade clinic infrastructure to accommodate VL services and ensure access to essential medical equipment and technology for effective treatment. |
|  | Change physical structure and equipment | 32 |  |
| **Communication** |  |  | Improve communication between healthcare workers, patients, and external partners to streamline service delivery and enhance information sharing. |
| **Culture** | Identify and prepare champions | 52 | Empower local opinion leaders and health champions to promote VL treatment, increasing community awareness and service uptake. |
| **Tension for change** | Identify and prepare champions | 48 | Identify key community leaders to champion change, foster consensus on VL treatment, and continuously assess local needs to adjust services accordingly. |
|  | Conduct local consensus discussions | 43 |  |
|  | Conduct local needs assessment | 43 |  |
|  | Inform local opinion leaders | 39 |  |
| **Incentive systems for healthcare providers to deliver VL services** | Alter incentive/allowance structures | 71 | Implement performance-based incentives for health workers delivering VL services to boost motivation and improve patient outcomes. |
|  | Access new funding | 38 |  |
| **Available Resources: Physical space, materials and equipment** | Access new funding | 78 | Secure funding to upgrade health facility infrastructure and procure necessary medical equipment for VL diagnostics and treatment. |
|  | Change physical structure and equipment | 48 |  |
|  | Fund and contract for clinical innovation | 39 |  |
| **Access to continuous training, updated knowledge & information** | Conduct educational meetings | 79 | Organize regular training sessions and workshops to update health workers on VL service guidelines. Develop and distribute educational materials tailored to local conditions. |
|  | Develop educational materials | 59 |  |
|  | Distribute educational materials | 55 |  |
|  | Create a learning collaborative | 45 |  |
|  | Conduct ongoing training | 38 |  |
| **High level leaders** |  |  | Engage high-level health leaders to advocate for increased resource allocation and policy support for VL services. |
| **Opinion Leaders** | Identify and prepare champions | 64 | Empower respected community figures to act as opinion leaders and VL service advocates, ensuring they spread awareness and promote service uptake. |
|  | Inform local opinion leaders | 57 |  |
|  | Build a coalition | 32 |  |
| **Role of service facilitators** |  |  | Train facilitators to coordinate service delivery, manage resources effectively, and support frontline workers. |
| **Planning** | Develop a formal implementation blueprint | 73 | Develop a structured VL implementation plan based on local assessments, clearly defining roles, timelines, and resource needs. |
|  | Conduct local needs assessment | 50 |  |
|  | Assess for readiness and identify barriers and facilitators | 42 |  |
| **Stakeholder engagement** | Identify and prepare champions | 63 | Actively engage patients, families, and community members in VL care through awareness campaigns and patient-centered education to ensure adherence to treatment. |
|  | Involve patients/consumers and family members | 59 |  |
|  | Prepare patients/consumers to be active participants | 55 |  |
|  | Intervene with patients/consumers to enhance uptake & adherence | 50 |  |
| **Reflecting & evaluating** | Develop and implement tools for quality monitoring | 60 | Establish systems for regular quality audits of VL services and provide feedback to health workers to ensure continuous service improvement. |
|  | Audit and provide feedback | 56 |  |
|  | Develop and organize quality monitoring systems | 40 |  |
